# Digital cognitive behavioural therapy for insomnia and primary care costs in England: an interrupted time series analysis

**DOI:** 10.1101/2021.02.15.21249646

**Authors:** Chris Sampson, Eleanor Bell, Amanda Cole, Christopher B. Miller, James Rose

**Affiliations:** Office of Health Economics, London, UK; Big Health Ltd., London, UK; Oxford AHSN, Oxford, UK

## Abstract

**Background:** *Sleepio* is an automated digital programme that delivers cognitive behavioural therapy for insomnia. *Sleepio* has been proven effective in improving sleep difficulties. However, evidence for the possible impact of *Sleepio* use on health care costs in the United Kingdom has not previously been developed. In this study, we assessed the effect of a population-wide rollout of *Sleepio* in terms of primary care costs in the National Health Service (NHS) in England.

**Methods:** The study was conducted in the Thames Valley region of England, where access to *Sleepio* was made freely available to all residents between October 2018 and January 2020. We use primary care data for people with relevant characteristics from nine general practices in Buckinghamshire. The study relies on a quasi-experimental design, using an interrupted time series to compare the trend in primary care costs before and after the rollout of *Sleepio*. Primary care costs include general practice contacts and prescriptions. Segmented regression analysis was used to estimate primary and secondary outcomes.

**Results:** For the 10,704 patients included in our sample, the total saving over the 65-week follow-up period was £71,027. This corresponds to £6.64 per person in our sample or around £70.44 per *Sleepio* user. Secondary analyses suggest that savings may be driven primarily by reductions in prescriptions.

**Conclusion:** Sleepio rollout reduced primary care costs. National adoption of *Sleepio* may reduce primary care costs by £20 million in the first year. The expected impact on primary care costs in any particular setting will depend on the uptake of *Sleepio*.

## Background

Poor sleep, sleep difficulty, or insomnia has a negative impact on people’s health-related quality of life [1]. In England, approximately 38% of individuals were found to have symptoms of insomnia, and the prevalence of diagnosed insomnia was 5.8% of the population in 2007 [2]. Insomnia often presents as a comorbid disorder alongside psychiatric and physical health conditions [3,4]. It is associated with an increased risk of developing other mental and physical health conditions, including anxiety, depression, alcohol misuse, psychosis [5], and cardiometabolic disease [6–8]. Insomnia is a public health concern, with the costs associated with insufficient sleep estimated to be around £40 billion in the UK due to lost productivity [9]. Further costs are thought to be generated from increased health care expenditure and accident risk [10], with higher health care costs for people co-presenting with insomnia and comorbidities [11].

Insomnia is most commonly managed in the UK by general practitioners through verbal advice (100%), minimally effective sleep hygiene education (89%), and sleep-promoting medication [12]. Patients rarely receive access to first-line cognitive behavioural therapy (CBT) for insomnia [4,13]. This is due in part to a shortage of trained providers [14] and a lack of treatment awareness [12,15]. CBT delivered by digital means offers a potential solution by facilitating population access because smartphones, tablets, and computers are now widely available. *Sleepio* is a standardised and fully automated (i.e., a standalone programme without the need for human input) digital therapeutic, which comprises the full range of cognitive and behavioural techniques used in CBT for insomnia [16]. It is designed for adults who want to improve their sleep and is available through a website and supporting iOS app. There is a growing body of evidence to support the effectiveness of *Sleepio*. For example, Espie et al. [17] and Freeman et al. [18] demonstrated benefit in terms of the Sleep Condition Indicator and Insomnia Severity Index, which capture insomnia symptoms including the time to fall asleep and night-time waking. Espie et al. [19] found benefits to functional health, well-being, sleep-related quality of life, and anxiety and depression symptoms.

The societal costs of insomnia are substantial [20], and evidence suggests that treatment for insomnia can be cost-saving [10]. *Sleepio* has previously been shown to improve workplace productivity [21,22]. Yet, its impact on health care service use and prescription costs is mostly unknown. To date, there has been some evidence published showing that use of *Sleepio* can be associated with reductions in self-reported medication use [23,24].

In England, *Sleepio* could be provided free-of-charge to the whole population, or to a subset of people who might need it, through National Health Service (NHS) funding. To guide such a decision, and to inform implementation strategies, real-world evidence can provide valuable insights beyond evidence generated in a trial setting.

In practice, *Sleepio* may be a substitute for alternative care strategies, increasing or decreasing health care costs. Substitutes may include ineffective management strategies based on (low cost) sleep-promoting medications [14], in which case *Sleepio* may be associated with better patient outcomes at higher costs. *Sleepio* might also be used as a substitute for more expensive management, such as regular general practitioner (GP) appointments. It may also be used where face-to-face behavioural therapy is not available [25]. Alternatively, *Sleepio* may complement other forms of health care and therefore increase health care costs. The impact of *Sleepio* is, *a priori*, ambiguous. To date, no evidence has been published to demonstrate the overall effect of *Sleepio* on primary care costs in practice.

This study sought to address this gap in knowledge by using real-world data to evaluate the impact of providing public access to *Sleepio*, in terms of i) primary care service use costs and ii) prescription costs. Our main objective was to identify whether providing access to *Sleepio* resulted in a change in the trend of total primary care costs from the perspective of the NHS in England.

## Methods

### Intervention

The *Sleepio* programme consists of six 20-minute digital CBT for insomnia sessions over at least six weeks, with each session unlocked after at least one week. People in therapy can maintain a sleep diary to track progress, with advice tailored to the provided information. The sleep diary can be automatically populated with data from a wearable device. Patients can also access other online support tools, including an electronic library and user community. *Sleepio* is currently priced per individual patient licence per year.

In this study, we do not conduct a cost-effectiveness analysis and do not consider any direct costs of *Sleepio*. This study evaluates population rollout of *Sleepio*, with primary care engagement, compared with current practice.

Population rollout involved all residents of the Thames Valley region of England (including Oxfordshire, Berkshire, and Buckinghamshire) being granted permission to access *Sleepio* free of charge. Eligibility was determined by the registrant’s postcode, which is entered when individuals first use *Sleepio*. Passive promotional activity, such as online and print advertising, was conducted throughout the region to encourage individuals to use *Sleepio*. Some employers in the region were also engaged and encouraged to promote *Sleepio* to their staff.

In Buckinghamshire, an additional strategy of primary care engagement was employed. This involved the *Sleepio* team working closely with selected general practices to offer *Sleepio* to patients most likely to benefit from digital CBT for insomnia. This involved training, implementing digital prompts for GPs, and providing patient-centred resources to each practice. During the rollout period, additional awareness material was distributed, and practices were given tailored support.

According to the employed implementation strategy, we refer to areas as either Tier 1 or Tier 2. Buckinghamshire, where primary care engagement was used, is Tier 1. Oxfordshire and Berkshire are areas that only received passive promotional activity and constitute Tier 2. The focus of this study is on Tier 1.

### Study design

This study adopts a before and after quasi-experimental design. Interrupted time series (ITS) analysis has been described as “the strongest, quasi-experimental approach for evaluating longitudinal effects of intervention” [26]. We used an ITS approach to estimate the change in total primary care costs following the rollout of *Sleepio*. The ITS analysis controlled for baseline levels and trends in costs.

The cohort was made up of patients from nine general practices within the primary care engagement (Tier 1) area. Patients that we expected to be more likely to use *Sleepio* (based on criteria described below) were selected to reduce noise in the sample.

Our study design makes two key assumptions. First, we assume that the primary care costs of people who do not use *Sleepio* are not affected by *Sleepio* rollout. Second, we assume that the people who do not satisfy our selection criteria do not use the *Sleepio* programme. These two assumptions imply that there would be no change in primary care use outside of our sample that is attributable to *Sleepio* rollout.

There may be a small proportion of people outside of our sample who use *Sleepio*, but we have no reason to expect that the direction of effect would be the opposite of that in our observed sample. Therefore, we expect these assumptions to provide conservative estimates.

### Data sources

#### EMIS data

EMIS Health is a software company that provides electronic patient record systems to general practices across the UK, including in Buckinghamshire. Nine GP practices in Buckinghamshire were recruited, from which we aimed to extract data for at least 10,000 patients.

Practices were selected such that the sample provides data for practices based in areas with a range of levels of deprivation. From each practice, patient-level data were extracted by an external company according to inclusion criteria. To be included in the extraction, patients needed to satisfy at least one of the following criteria within the extraction period:

1. A diagnosis of insomnia
2. A diagnosis of depression or anxiety disorder
3. Prescription of a hypnotic or anxiolytic medication
4. Referral to *Sleepio* by a GP

Individuals below the age of 18 years at the time of data extraction were excluded from the data set. The purpose of these criteria is to limit our sample to those individuals that we would anticipate might use (and potentially be affected by the rollout of) *Sleepio*. We expected that insomnia diagnosis would rarely be coded in the data. Therefore, relevant prescribing (i.e., BNF chapter 4, section 1 drugs) was used as an inclusion criterion to identify people experiencing sleep problems.

Table 9 in the Appendix lists the data items that were extracted from EMIS. Data were aggregated as patient-weeks, except for time-invariant patient characteristics. Weekly observations were judged to provide a sufficient number of data points to conduct the regression analyses described below and allow adequate precision in identifying exposure to *Sleepio*.

The data extraction period was 12 months before *Sleepio* rollout (October 2017), up to 15 months after *Sleepio* rollout (January 2020). This provided an adequate timeframe to capture seasonal effects within the ITS design. Notably, our timeframe incorporates three Christmas periods, which we anticipated would be a significant correlate for primary care service use. Service use tends to be lower around Christmas, creating an artificial downward trend in primary care costs if not adequately controlled. Data were extracted by an independent specialist provider (Interface Clinical Services Ltd).

#### Sleepio data

Data were collected from all individual users of *Sleepio*, either through the *Sleepio* website or the *Sleepio* iOS app. This sample cannot be linked to the EMIS sample. These data are used for descriptive purposes only, providing information on uptake across the region.

### Analysis

#### Primary analysis

The primary outcome for the analysis is the average primary care costs per patient per week, where primary care costs include GP practice contacts and prescription costs. Unit costs were attributed to resource use using information from the Unit Costs of Health and Social Care [27]. According to the specific medicine, dose, and pack size, prescription costs were obtained from the BNF Online [28].

We employed a segmented regression analysis of the interrupted time series data to estimate the change in the trend of the primary outcome, such that the full model was estimated as

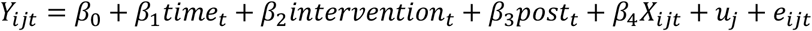

where *Y*_*ijt*_ is total primary care costs for individual *i* from practice *j* at time *t*; *time*_*t*_ corresponds with the number of the week in the time series at time *t*; *intervention*_*t*_ is a binary indicator for time *t* being before (*intervention* = *0*) or after (*intervention* = 1) *Sleepio* rollout; *post*_*t*_ corresponds to the number of weeks after the intervention at time *t*; *X*_*ijt*_ is a vector of patient-, practice-, and time-specific confounders; *u*_*j*_ represents a random error term at the level of the practice; and *e*_*ijt*_ represents unexplained variability.

In this model, *β*_*0*_ estimates baseline costs at *t* = *0*; *β*_1_ estimates the pre-*Sleepio* trend in costs; *β*_2_ estimates the immediate change in *Y* at the time of *Sleepio* rollout; and *β*_3_ estimates the change in trend after *Sleepio* rollout compared with the pre-intervention trend. The post-intervention slope is estimated as *β*_1_ + *β*_3_. Seasonal effects are accounted for by including an indicator within *X* for the season in which week *t* falls, where weeks 10-22 of any calendar year are spring, 23-35 are summer, 36-48 are autumn, and 49 through to 9 are winter. *β*_4_ estimates seasonal and other confounding effects. We assume the rollout period for *Sleepio* to span six weeks.

The segmented regression analysis used a generalised linear model, with appropriate distributions and link functions fitted according to visual inspection of the data and use of a modified Park test and link tests. First-order autocorrelation was tested using the Durban-Watson test.

Individual-level observations were drawn from practices, within which observations may be correlated. We therefore implemented a multilevel regression model to account for clustering within practices.

The ITS approach’s key underlying assumption is that the pre-*Sleepio* trend in primary care costs for this group of patients would have continued had *Sleepio* not been introduced. Thus, we assume that no other interventions, policy changes, or other external factors affect the trends. Another critical assumption is that pre-intervention trends are linear. We tested this assumption in interim and final analyses through visual inspection.

#### Secondary analyses

We conducted four exploratory secondary analyses, as set out in Table 1: Overview of secondary analyses. We anticipated that diagnoses of insomnia would be poorly recorded and that the sample would be relatively small. Therefore, we did not plan to conduct secondary analysis on this subgroup.

**TABLE 1:**
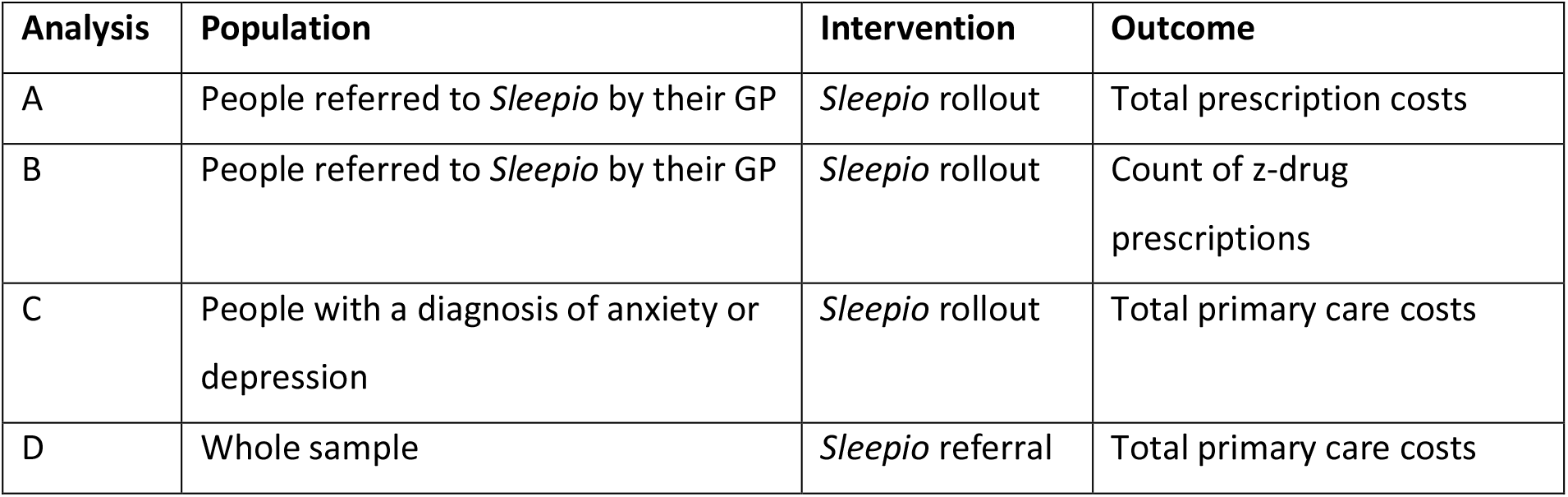
OVERVIEW OF SECONDARY ANALYSES.

Two of our secondary analyses (A and B) focussed on prescriptions. These analyses could help health care commissioners to better understand the impact on costs and resource use characterised in our primary analysis. The budgetary implications of changes in prescription costs may be different from those associated with changes in GP contacts. The long-term prescription of nonbenzodiazepines (zolpidem and zopiclone, commonly known as z-drugs) for sleep problems is a concern in itself, due to their side effects and lack of effectiveness [13].

For these analyses (A and B), we only included individuals who were – at any point in our follow-up period – referred to *Sleepio*. Reducing the size of the dataset in this way made it possible to more effectively control for individual-level variation and potentially identify effects directly attributable to *Sleepio* use.

Previous research on *Sleepio* has demonstrated benefits in terms of anxiety or depression symptoms, and the potential for *Sleepio* to be particularly effective for people with anxiety or depression [18,19,29]. To contribute to this evidence base, we implemented our primary analysis with stratification according to the presence of a diagnosis of anxiety or depression. In this secondary analysis (C), the model was run separately for the two groups (with and without a diagnosis) and the findings compared. This analysis facilitates predictions about the cost implications of providing access to *Sleepio* for people with anxiety or depression, compared with the broader population.

To further test the robustness of our findings and the extent to which any treatment effect is attributable to *Sleepio* rollout, we conducted an analysis (D) whereby referral to *Sleepio* is the intervention. In this case, the control group is the population of patients who were never referred to *Sleepio*. We implemented an analysis similar to our primary analysis, using a hierarchical generalised linear model to evaluate the effect on primary care costs, in any given week, of having been referred to *Sleepio* by a GP. This approach is more susceptible to selection bias but could provide more precise identification of *Sleepio* patients and reduce noise in the sample.

## Results

### Descriptive statistics

The EMIS data set included 1,252,368 person-week observations from 10,704 people over 117 weeks from October 2017 to January 2020. 64.41% of the sample identified as female. Table 2 shows the number of people in each practice and the number recorded as being referred to *Sleepio* at least once within each practice, of which there were 1,008 people in total.

**TABLE 2:**
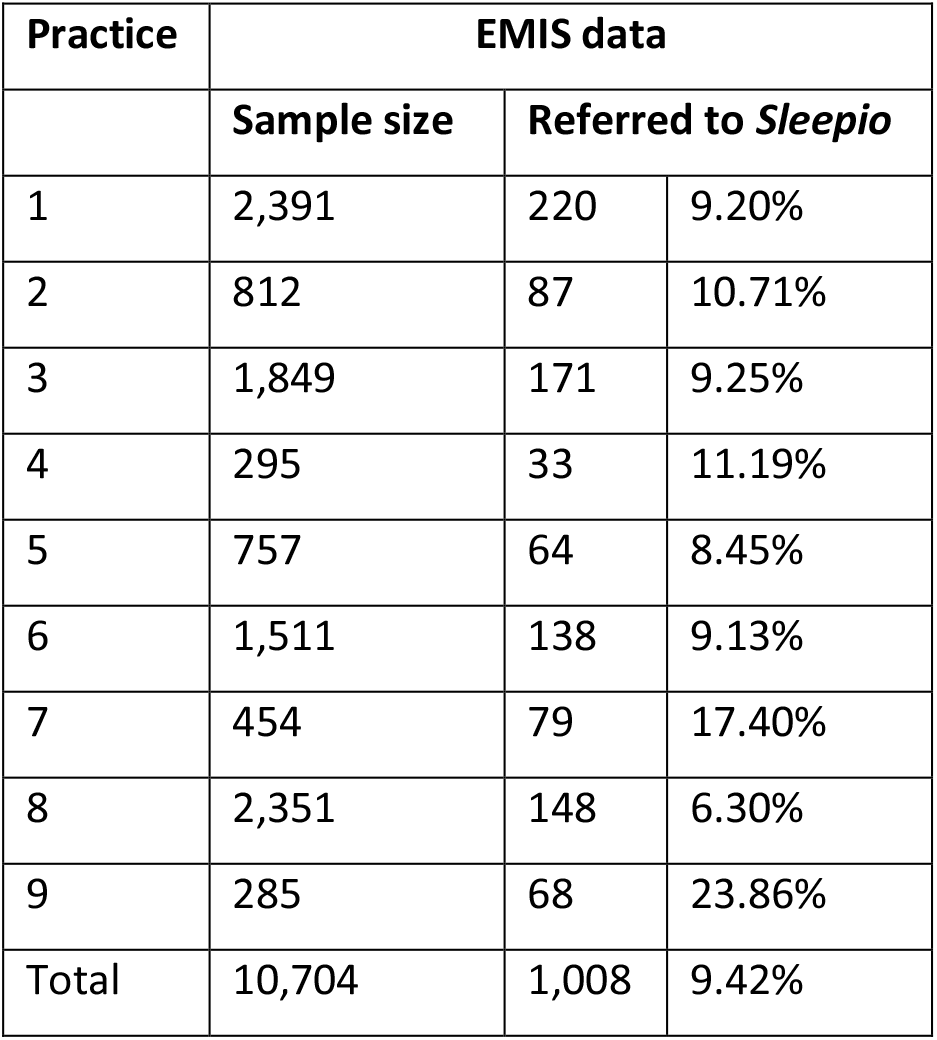
GP PRACTICE SAMPLES.

Across the sample, 3,001 people received at least one of the two types of inclusion criteria prescriptions (hypnotics or anxiolytics), with 1,919 receiving at least one hypnotic prescription and 1,502 receiving at least one anxiolytic prescription.

In October 2018, the total number of patients registered with the nine practices in our study was 129,865, and the total population of Buckinghamshire was around 540,059 [30]. Table 3 shows the estimated number of *Sleepio* patients for different population sizes, based on the number of people recorded as being referred by their GP in the EMIS data and the actual number of patients recorded by *Sleepio*, within the full 117 weeks of the study.

**TABLE 3:**
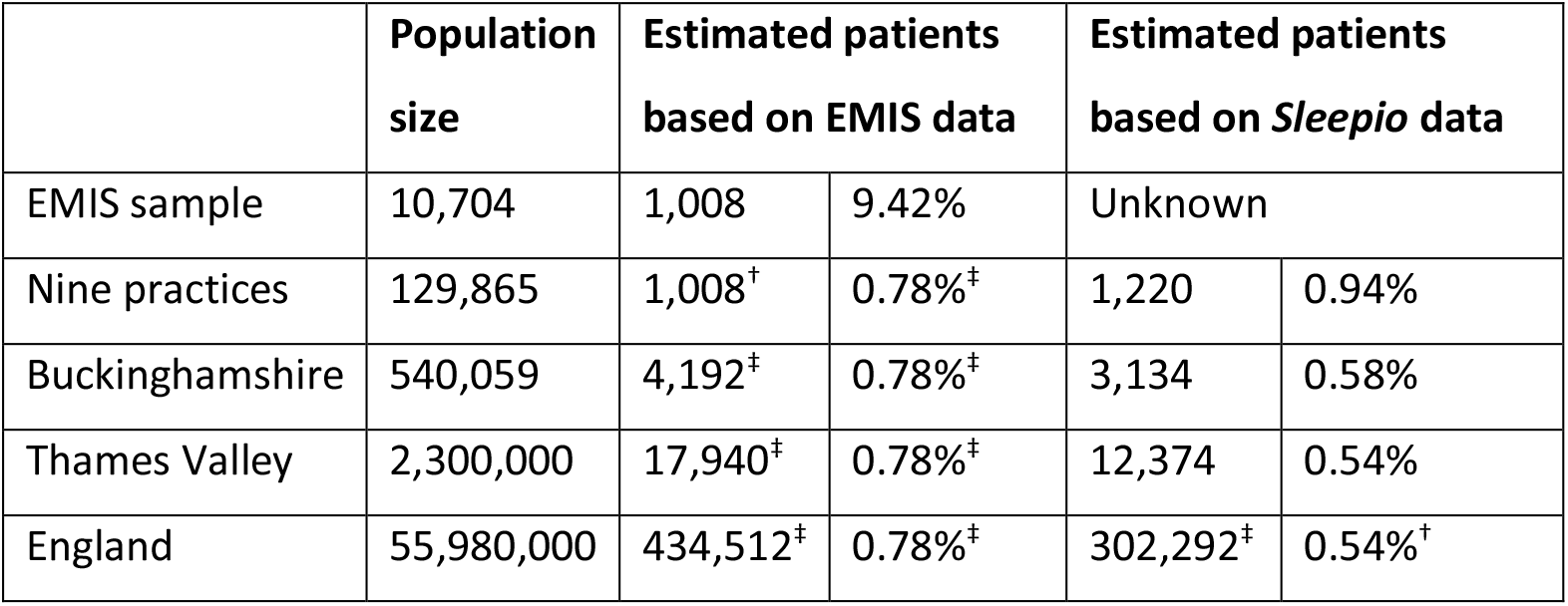
ESTIMATED NUMBER OF SLEEPIO PATIENTS BY POPULATION (^†^EXTRAPOLATED FROM OBSERVATION; ^‡^ASSUMED FROM EXTRAPOLATION)

The EMIS data only included referrals recorded by GPs, while the *Sleepio* data included all referral routes. *Sleepio* data relating to the nine practices rely on patients self-reporting GP referral. An individual’s county is determined by their postcode provided in *Sleepio*. We assume the Thames Valley region to be formed of the three counties of Buckinghamshire (including Milton Keynes), Oxfordshire, and Berkshire.

Figure 1 shows the average primary care costs per person per week in the whole sample, with general practice contacts (including GP and nurse face-to-face contacts and telephone contacts) and prescription costs separately. The black line shows the time trend before, during, and after the six-week rollout period for *Sleepio*, with 95% confidence intervals for the trends derived by linear regression.

**FIGURE 1:**
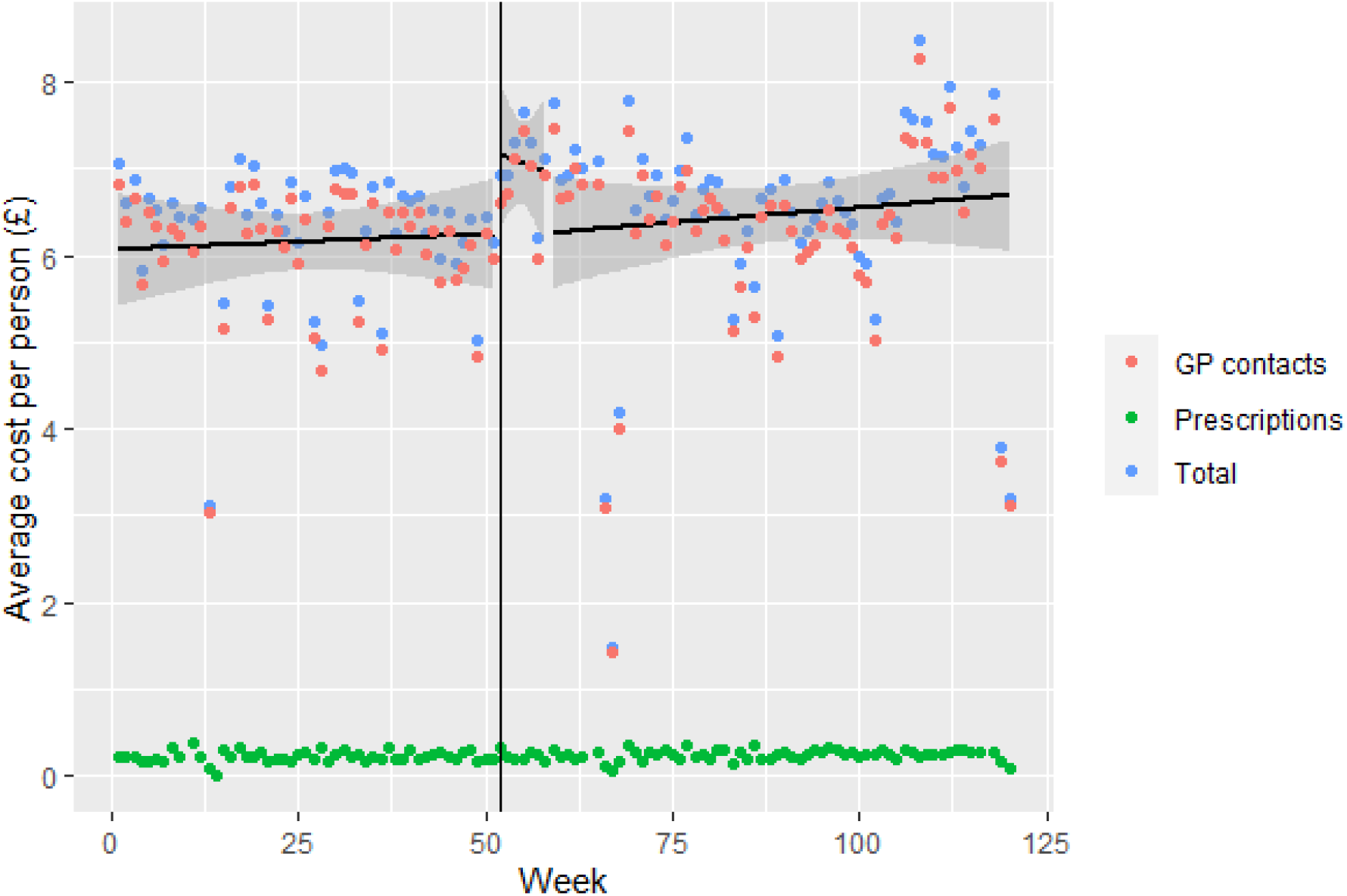
AVERAGE COSTS (£) PER PERSON PER WEEK.

Figure 1 suggests that there is no clear change in trend in costs associated with *Sleepio* rollout. However, the trend is likely subject to seasonal effects and other confounding factors that may obscure any underlying trend.

Table 4 shows the average cost per person per week in the pre-*Sleepio* period and the post-*Sleepio* period for various groups. Almost all groups show an increase in costs in the post-*Sleepio* period, reflecting the increasing trend in primary care costs across the sample.

**TABLE 4:** AVERAGE PRIMARY CARE COSTS PER PERSON PER WEEK.

|  | Sample size |  | Pre-Sleepio |  | Post-Sleepio |  |
| --- | --- | --- | --- | --- | --- | --- |
|  | # | % | Mean | Std Dev | Mean | Std Dev |
| Whole sample | 10,704 | 100% | £6.16 | 19.6 | £6.53 | 19.9 |
| Referred to *Sleepio* | 1,008 | 9% | £5.77 | 18.7 | £6.90 | 19.9 |
| *Practice* |  |  |  |  |  |  |
| 1 | 2,391 | 22% | £5.71 | 17.7 | £5.91 | 17.9 |
| 2 | 812 | 8% | £5.64 | 17.7 | £5.38 | 16.8 |
| 3 | 1,849 | 17% | £6.72 | 20.8 | £6.31 | 19.3 |
| 4 | 295 | 3% | £8.03 | 24.3 | £9.04 | 25.3 |
| 5 | 757 | 7% | £7.18 | 22.3 | £8.22 | 22.7 |
| 6 | 1,511 | 14% | £7.03 | 21.4 | £7.93 | 22.5 |
| 7 | 454 | 4% | £4.80 | 14.6 | £5.23 | 15.1 |
| 8 | 2,350 | 22% | £6.20 | 20.3 | £6.99 | 21.4 |
| 9 | 285 | 3% | £0.21 | 1.98 | £0.29 | 3.89 |
| *Diagnoses* |  |  |  |  |  |  |
| Anxiety or depression | 5,515 | 52% | £5.78 | 18.7 | £6.03 | 18.8 |
| Arthritis | 98 | 1% | £10.00 | 23.6 | £10.90 | 24.6 |
| Asthma | 835 | 8% | £8.95 | 23.8 | £9.37 | 24.0 |
| COPD | 313 | 3% | £11.00 | 26.1 | £11.80 | 26.3 |
| Diabetes | 739 | 7% | £9.73 | 24.4 | £10.10 | 24.2 |
| Heart failure | 117 | 1% | £12.50 | 28.0 | £13.90 | 28.4 |
| Hypertension | 1,322 | 12% | £7.72 | 21.2 | £7.89 | 21.1 |
| IHD | 296 | 3% | £10.20 | 24.5 | £10.30 | 24.2 |
| Insomnia | 1,655 | 15% | £6.30 | 20.0 | £6.61 | 19.9 |
| Chronic pain | 2,959 | 28% | £8.28 | 22.6 | £8.68 | 22.9 |
| Any diagnosis (of the above) | 8,146 | 76% | £6.25 | 19.5 | £6.54 | 19.7 |
| No diagnosis (of the above) | 2,558 | 24% | £5.86 | 20.0 | £6.50 | 20.6 |

### Primary analysis

The regression results from the primary analysis are shown in Table 5. We report several models as a sensitivity analysis of alternative specifications. For instance, our models test the sensitivity of assumptions about seasonal adjustment and assuming fixed effects for different diagnoses.

**TABLE 5:** FULL SAMPLE REGRESSION COEFFICIENTS (SIGNIFICANCE LEVELS: *** < 0.001; ** < 0.01; * < 0.05)

|  | **Preferred model** | **Model 2** | **Model 3** | **Model 4** | **Model 5** |
| --- | --- | --- | --- | --- | --- |
| Time | 0.002*** | 0.002*** | 0.002*** | 0.001*** | 0.004 |
| Intervention | -0.038*** | -0.036*** | -0.033*** | 0.033*** | 0.263*** |
| Post | -0.002*** | -0.002*** | -0.002*** | -0.000 | -0.003 |
| Seasonal adjustment | YES | YES | YES | NO | NO |
| Age bands | YES | YES | NO | NO | NO |
| Gender | YES | YES | NO | NO | NO |
| Diagnoses | YES | NO | NO | NO | NO |
| Practice random effects | YES | YES | YES | YES | NO |

The preferred model and models 2 to 4 are hierarchical generalised linear models using a quasi-gamma distribution with a variance function of *V(μ)*= μ^2^ and log link. Model 5 is an ordinary least squares linear regression model. Our preferred model is that which exhibited superiority in statistical tests and provided the most convincing predictions visually.

We focus on a description of the findings from the preferred model, presenting the alternative models’ results to demonstrate the robustness of our findings to alternative specifications.

A positive coefficient for *Time* shows that primary care costs increase over time, prior to *Sleepio* rollout. A negative coefficient for *Intervention* implies that the immediate impact of *Sleepio* rollout (during the six-week rollout period) is to reduce primary care costs. A negative coefficient for *Post* shows that the effect of *Sleepio* rollout was to reduce the trend shown by the coefficient on *Time*. If the negative coefficient for *Intervention* is smaller than the positive coefficient for *Time*, it implies that the upward trend in primary care costs continues after *Sleepio* rollout, but at a slower rate.

The regression results imply three key overall findings. First, there is a small increasing trend in primary care costs over time, as illustrated in Figure 1. Second, *Sleepio* rollout has a small but statistically significant negative impact on costs during the initial six-week rollout period (contrary to the naïve indication in Figure 1, quantified in Model 5). Third, *Sleepio* rollout mitigates the trend of increasing primary care costs.

Comparison between the alternative model specification reveals that seasonal adjustment is critical; accounting for seasonal effects reverses the direction of effect for *Intervention* and results in statistical significance for *Post*.

Our preferred model results show that the absolute difference in mean weekly costs per person, associated with *Sleepio* rollout, is a saving of £0.16 at week 65. This corresponds to £6.64 per person over the 65-week follow-up period, including the initial rollout period. The 95% confidence interval for this estimate is a saving of between £4.60 and £8.67. Across the observed sample of 10,704 people, *Sleepio* rollout reduced primary care costs by £71,027 (95% confidence interval £49,291 to £92,762).

Table 6 presents projections for average cost savings for different populations over different durations, based on our preferred model specification. The projections assume that, beyond our observation period (i.e., 65 weeks), the trend in primary care costs returns to the trend observed before *Sleepio* rollout (represented by the coefficient for *Time* in Table 5).

**TABLE 6:** PROJECTED REDUCTION IN PRIMARY CARE COSTS ASSOCIATED WITH SLEEPIO ROLLOUT (^†^PROJECTION BEYOND THE OBSERVED PERIOD; *9.42% OF OUR SAMPLE BASED ON GP REFERRALS; CI: CONFIDENCE INTERVAL)

| Sample | 1 year | 65 weeks | 2 years<sup>†</sup> | 3 years<sup>†</sup> |
| --- | --- | --- | --- | --- |
| Per person<br>(95% CI) | £4.66<br>(£3.20 – £6.13) | £6.64<br>(£4.60 – £8.67) | £13.00<br>(£9.15 – £16.84) | £21.48<br>(£15.22 – £27.75) |
| Per *Sleepio* user* | £49.52 | £70.44 | £138.00 | £228.07 |
| Nine practices | £49,930 | £71,027 | £139,144 | £229,967 |
| Buckinghamshire | £207,640 | £295,374 | £578,648 | £956,347 |
| Thames Valley | £884,298 | £1,257,936 | £2,464,343 | £4,072,885 |
| England | £21,523,042 | £30,617,071 | £59,979,956 | £99,130,468 |

Table 6 includes estimates per *Sleepio* user, based on the uptake estimates in Table 3, which assumes a growing population of users with growth at the rate observed in our study. Economic evaluations and decision analyses often rely on the estimation of effects at the individual level, which our study does not observe. Therefore, to inform future research, we provide alternative projections based on changes over time at the individual level. These projections are based on the assumption that *Sleepio* rollout is equivalent to treatment exposure at the individual level, such that those people identified as *Sleepio* users become *Sleepio* users at the point of the rollout. These projections assume a return to pre-rollout trends and no new users after year 1, such that year 1 savings for projected new users are subtracted from projected cumulative savings. In this case, the two-year savings per user would be £88.48 (£138.00 *minus* £49.52). The three-year savings would be £139.59 (£228.07 *minus* £88.48). Rather than projecting a growing saving over time, as trends diverge, this approach assumes a shift to the pre-rollout trend to converge on an annual saving of £90.07 per user in the long-term. Based on existing evidence, these effects might be expected to be maintained for up to three years [31].

### Secondary analyses

Table 7 shows the key coefficients for our secondary analyses that maintained the segmented regression analysis approach, as summarised in Table 1. All analyses included seasonal adjustment, age, gender, diagnoses, and practice random effects. Analysis A and Analysis B used a Poisson distribution with a log link. Analysis C used the same model specification as our primary analysis.

**TABLE 7:**
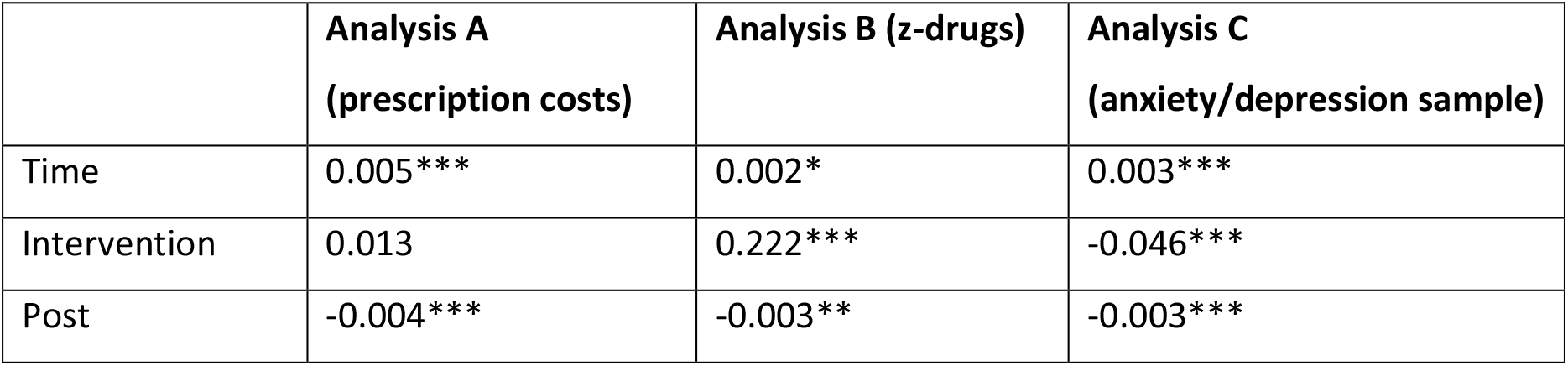
SECONDARY ANALYSIS REGRESSION COEFFICIENTS.

The coefficient for the pre-existing increasing trend in prescription costs (0.005) is greater than that for overall costs (0.002), as is the coefficient for the impact of *Sleepio* on the trend (−0.004 compared with −0.002). Analysis A shows that the reduction in prescription costs at 65-week follow-up is £8.62 per person (£5.40 in the first year), suggesting that our primary analysis’s observed reductions in prescription costs may significantly explain savings. Analysis B shows that *Sleepio* had a small but statistically significant impact in achieving a downward trend in the prescription of z-drugs. Analysis C supports the notion that *Sleepio* may be more effective in reducing costs among people diagnosed with anxiety or depression. The average saving per person in this group, over 65 weeks, was £9.27.

Our final analysis (D), treating *Sleepio* referral as the intervention, identified a negative but statistically insignificant effect. This is likely due to a selection bias, whereby individuals who use *Sleepio* may be more likely to have higher levels of resource use, all else equal. An exploratory analysis of the insomnia subsample did not identify any statistically significant findings.

Table 8 shows the total costs and counts associated with the ten most-prescribed medicines in the subset of 2,122 person-week observations in which a diagnosis of insomnia was recorded. These data demonstrate that z-drugs are commonly prescribed to people presenting in primary care with insomnia. Due to these drugs’ low unit cost, it is unlikely that a reduction in z-drug prescribing for people with insomnia explains the significant cost reduction observed.

**TABLE 8:** MEDICINES COMMONLY PRESCRIBED FOR PEOPLE WITH INSOMNIA (*CORRESPONDS TO PACK SIZE, SUCH AS NUMBER OF TABLETS)

| Drug | Count* | Cost |
| --- | --- | --- |
| Zopiclone 3.75mg tablets | 8315 | £317.75 |
| Amitriptyline 10mg tablets | 5313 | £191.65 |
| Zopiclone 7.5mg tablets | 2911 | £112.28 |
| Temazepam 10mg tablets | 1262 | £77.52 |
| Diazepam 2mg tablets | 399 | £10.12 |
| Diazepam 5mg tablets | 299 | £8.12 |
| Zolpidem 10mg tablets | 276 | £13.60 |
| Amitriptyline 50mg tablets | 252 | £20.52 |
| Amitriptyline 25mg tablets | 238 | £7.40 |
| Zolpidem 5mg tablets | 140 | £5.40 |

**TABLE 9:** EMIS DATA ITEMS.

| Variable | Type | Description |
| --- | --- | --- |
| **ID** | String | Patient identifier (not NHS number), fixed |
| **Practice_ID** | String | Anonymised practice identifier, fixed |
| **Year** | Numeric | Year |
| **Week.Number** | Numeric | Week of Year (0-52) |
| **Age_Band** | String | Categorised by five-year brackets, fixed |
| **Gender** | String | Female/male/other, fixed |
| **GP.Practice.Consultation** | Numeric | Number of GP practice consultations |
| **GP.Home.Consultation** | Numeric | Number of GP home consultations |
| **GP.Telephone.Consultation** | Numeric | Number of GP telephone consultations |
| **Nurse.Consultation** | Numeric | Number of nurse consultations |
| **Hospital.Referrals** | Numeric | Number of hospital referrals |
| **IAPT.Referrals** | Numeric | Number of referrals to IAPT |
| **CBT.Referrals** | Numeric | Number of referrals to CBT |
| **Sleep.Clinic.Referrals** | Numeric | Number of referrals to sleep clinic |
| **Sleepio.Referral** | Binary | Sleepio referral (yes/no) |
| **Insomnia.Diagnosis** | Numeric | Number of times a diagnosis of insomnia is recorded |
| **Anxiety.or.Depression.Diagnosis** | Numeric | Number of times a diagnosis of either anxiety or depression is recorded |
| **Diabetes.Diagnosis** | Numeric | Number of times a diagnosis of diabetes is recorded |
| **Hypertension.Diagnosis** | Numeric | Number of times a diagnosis of hypertension is recorded |
| **COPD.Diagnosis** | Numeric | Number of times a diagnosis of COPD is recorded |
| **Asthma.Diagnosis** | Numeric | Number of times a diagnosis of asthma is recorded |
| **IHD.Diagnosis** | Numeric | Number of times a diagnosis of ischaemic heart disease is recorded |
| **HF.Diagnosis** | Numeric | Number of times a diagnosis of heart failure is recorded |
| **Arthritis.Diagnosis** | Numeric | Number of times a diagnosis of arthritis is recorded |
| **Chronic.Pain.Diagnosis** | Numeric | Number of times a diagnosis of chronic pain is recorded |
| **Hypnotics.[...]** | Numeric | Quantity prescribed [for each dosage of each medicine] |
| **Anxiolytics.[...]** | Numeric | Quantity prescribed [for each dosage of each medicine] |
| **Barbiturates.[...]** | Numeric | Quantity prescribed [for each dosage of each medicine] |
| **Benzodiazepines.[...]** | Numeric | Quantity prescribed [for each dosage of each medicine] |
| **Amitriptyline.[...]** | Numeric | Quantity prescribed [for each dosage of each medicine] |
| **OtherMeds.[...]** | Numeric | Quantity prescribed [for each dosage of each medicine] |

## Discussion

Our analysis’s main findings show that the rollout of *Sleepio* in the Thames Valley region reduced average primary care costs, including general practice contacts and prescriptions.

Over the observed follow-up period, the average saving in our sample was £6.64 per person. Assuming that people outside of our sample were not affected by the rollout of *Sleepio*, this corresponds to a saving of £71,027 across the nine practices.

Our secondary analyses suggest that reductions in prescription costs are a significant driver in reducing overall primary care costs. This is partly explained by reductions in the prescription of z-drugs. However, off-label prescribing may explain cost savings as opposed to changes to on label z-drugs. Data from Sleepio suggest that around 26% of people referred to Sleepio would start CBT. Therefore, based on this analysis, the saving per patient associated with prescription costs may be £20 in the first year (£5.40 x 0.26%); around half of the per-user saving identified across the whole sample.

The reduction in costs observed in the subsample of people with a diagnosis of anxiety or depression was greater than the average saving across the whole sample. Future research should explore the potential for cost savings in different categories of expenditure and different populations.

A strength of our analysis is that the primary and secondary outcomes and analytical approach were determined before the data were analysed. The preferred model was selected on the basis of predictive ability and fit for the primary outcome.

Our main findings are robust across a variety of model specifications. The observed direction of effect on the trend in costs is not sensitive to seasonal effects or individuals’ characteristics. This supports the generalisability of our findings. Practices with different proportions of people with the diagnoses listed in Table 4 might, therefore, expect to observe similar impacts for the subpopulation that satisfies our inclusion criteria.

The inclusion of seasonal effects and practice random effects was important in demonstrating a statistically significant impact of *Sleepio* rollout. However, the direction of effect for the trend in costs was not undermined by their exclusion. Our evidence suggests that the cost savings were not driven by any single practice and this therefore supports the generalisability of our findings. However, we did observe variation by practice, and commissioners should consider the characteristics of providers and patients that might act as barriers or facilitators to changes in service use.

### Limitations

It is important to note that our study is not a cost-effectiveness analysis, and our estimates do not include other potential cost impacts associated with *Sleepio* rollout. In particular, our analysis does not attribute any direct cost to *Sleepio* rollout or use. The advantage of this is that our analysis is independent of *Sleepio* pricing, and the relevance of our study will not be undermined by changes in the price of *Sleepio* licences.

In practice, access to *Sleepio* may be priced on the basis of initiation of digital CBT for insomnia, rather than initial registration by a user or referral by a GP. A limitation of our analysis is that we cannot identify which individuals in our EMIS data sample initiated digital CBT for insomnia^1^.

We are also unable to observe any changes in resource use in Tier 2 areas, where primary care engagement was based on passive promotional activity and not used to drive uptake. As shown in Table 3, a lower level of uptake was observed in Tier 2 areas. This has implications for implementation strategies and related costs. Further work could evaluate the resource impact of *Sleepio* rollout in alternative settings, such as when delivered as an adjunctive intervention for those with poor sleep through the Improving Access to Psychological Therapies (IAPT) programme.

Our analysis does not provide estimates of any impact on referrals to secondary care or resources used in settings other than GP practices. Due to the pre-specification of our primary outcome, and the development of our model to suit these data, there was limited scope for us to explore different aspects of resource use without suffering from over-testing.

Optimal sample specifications for identifying statistically significant effects in interrupted time series analyses are difficult to estimate reliably [32]. One limitation of our study is that we did not conduct simulations before commencing data collection to specify a sample size. The sample size was determined on the basis of practicality and our expectations about uptake and the variability in health care costs. Nevertheless, the total number of observations is likely to provide reliable estimates [26].

## Conclusion

The rollout of *Sleepio* in the Thames Valley resulted in lower primary care costs across nine practices with one year. Providing NHS patients in England with access to *Sleepio*, while encouraging GPs to refer patients with sleep problems to register for *Sleepio*, is likely to result in fewer GP attendances and fewer prescriptions in the population. Therefore, direct costs associated with the adoption of Sleepio will be partially, or entirely, offset by these savings.

## Data Availability

The study used no identifiable information. As per the study's data sharing agreements, the data cannot be made available to other researchers.

## Ethics and data availability

This study involved no randomisation, did not require changes to accepted treatment standards within the NHS, and did not involve primary data collection. The study was retrospectively reviewed by the Joint Research Office study classification group at Oxford University Hospitals NHS Foundation Trust and classified as service evaluation, such that it did not require research ethics board approval. The study received permission from Buckinghamshire Clinical Commissioning Group’s Medicines Management Assurance Committee.

All Sleepio participants agreed to a privacy policy when they registered for Sleepio and consented to Big Health Ltd. using health information for research purposes. This allows for non-identifiable health information only to be published in aggregate form for academic research. Collection and analysis of data within the Sleepio programme was approved by the Medical Sciences Interdivisional Research Ethics Committee, University of Oxford (Ref R72295/RE001)

The study used no identifiable information. As per the study’s data-sharing agreements, the data cannot be made available to other researchers.

## Acknowledgements and disclaimers

Earlier versions of this analysis were shared with stakeholders and presented at academic meetings. We are grateful for comments received at the winter 2020 meeting of the Health Economists’ Study Group and ISPOR Europe 2019. The reported results in earlier versions may differ following improvements in the analysis.

## Footnotes

1 26% according to Big Health operational data from the Thames Valley experiment.

